# Receiving information on machine learning-based clinical decision support systems in psychiatric services increases staff trust in these systems: A randomized survey experiment

**DOI:** 10.1101/2024.09.09.24313303

**Authors:** Erik Perfalk, Martin Bernstorff, Andreas Aalkjær Danielsen, Søren Dinesen Østergaard

**Author notes:** **Corresponding author** Erik Perfalk, MD, Department of Affective Disorders Aarhus University Hospital - Psychiatry Palle Juul-Jensens Boulevard 175, 8200 Aarhus N Denmark.

## Abstract

**Background:** Clinical decision support systems based on machine learning (ML) models are emerging within psychiatry. To ensure their successful implementation, healthcare staff needs to trust these systems. Here, we investigated if providing staff with basic information about ML-based clinical decision support systems enhances their trust in them.

**Methods:** We conducted a randomised survey experiment among staff in the Psychiatric Services of the Central Denmark Region. The participants were allocated to one of three arms, receiving different types of information: An intervention arm (receiving information on clinical decision-making supported by an ML model); an active control arm (receiving information on standard clinical decision process without ML support); and a blank control arm (no information). Subsequently, participants responded to various questions regarding their trust/distrust in ML-based clinical decision support systems. The effect of the intervention was assessed by pairwise comparisons between all randomization arms on sum scores of trust and distrust.

**Findings:** Among 2,838 invitees, 780 completed the survey experiment. The intervention enhanced trust and diminished distrust in ML-based clinical decision support systems compared with the active control arm (Trust: mean difference= 5% [95% confidence interval (CI): 2%; 9%], p-value < 0.001; Distrust: mean difference=-4% [-7%; -1%], p-value = 0.042)) and the blank control arm (Trust: mean difference= 5% [2%; 11%], p-value = 0.003; Distrust: mean difference= -3% [-6%; - 1%], p-value = 0.021).

**Interpretation:** Providing information on ML-based clinical decision support systems in hospital psychiatry may increase healthcare staff trust in such systems.

## Introduction

Machine learning (ML) is rooted in the idea that machines (computers) may learn from historical data and be trained for pattern recognition (e.g., prediction). The benefits of using ML to aid decision-making in the medical field are promising.^1^ Indeed, prediction models based on ML have been shown to be accurate across medical fields, including psychiatry, achieving performance levels comparable to or exceeding those of clinicians.^2^ However, in psychiatry, only 0.4% of ML-based prediction models have been considered for implementation in clinical practice.^3^

To ensure successful implementation of ML prediction models into clinical contexts, it is essential that healthcare staff accept and trust this technology. Staff trust in ML models has previously been explored. These studies have generally found that staff believe that ML-based prediction models should not automatize clinical decisions, but rather serve as a clinical decision support systems, where the final clinical decision should rest with the staff.^4^ Additionally, information on how the model works (transparency) and the ability to explain clinical predictions (explainability) were considered as essential to establish trust.^4^

To our knowledge, no studies have investigated if staff members’ trust in ML-based clinical decision support systems can be increased by providing basic information on these tools. In a randomized survey experiment, we have previously demonstrated that receiving brief information regarding ML models as clinical decision support systems increased the trust in such systems among patients with mental disorders.^5^ It would seem likely that this intervention could have the same positive effect among staff working in psychiatric services. Therefore, this study explored that hypothesis in an analogue randomized survey experiment.

## Methods

### Design

In keeping with our analogue study among patients from the Psychiatric Services in the Central Denmark Region,^5^we conducted a randomized survey experiment to investigate whether receiving information about ML-based clinical decision support systems could increase staff trust in this technology. The study design and analysis plan were pre-registered and are available at https://doi.org/10.17605/OSF.IO/GST8X. The study design is illustrated in Figure 1.

**Figure 1.**
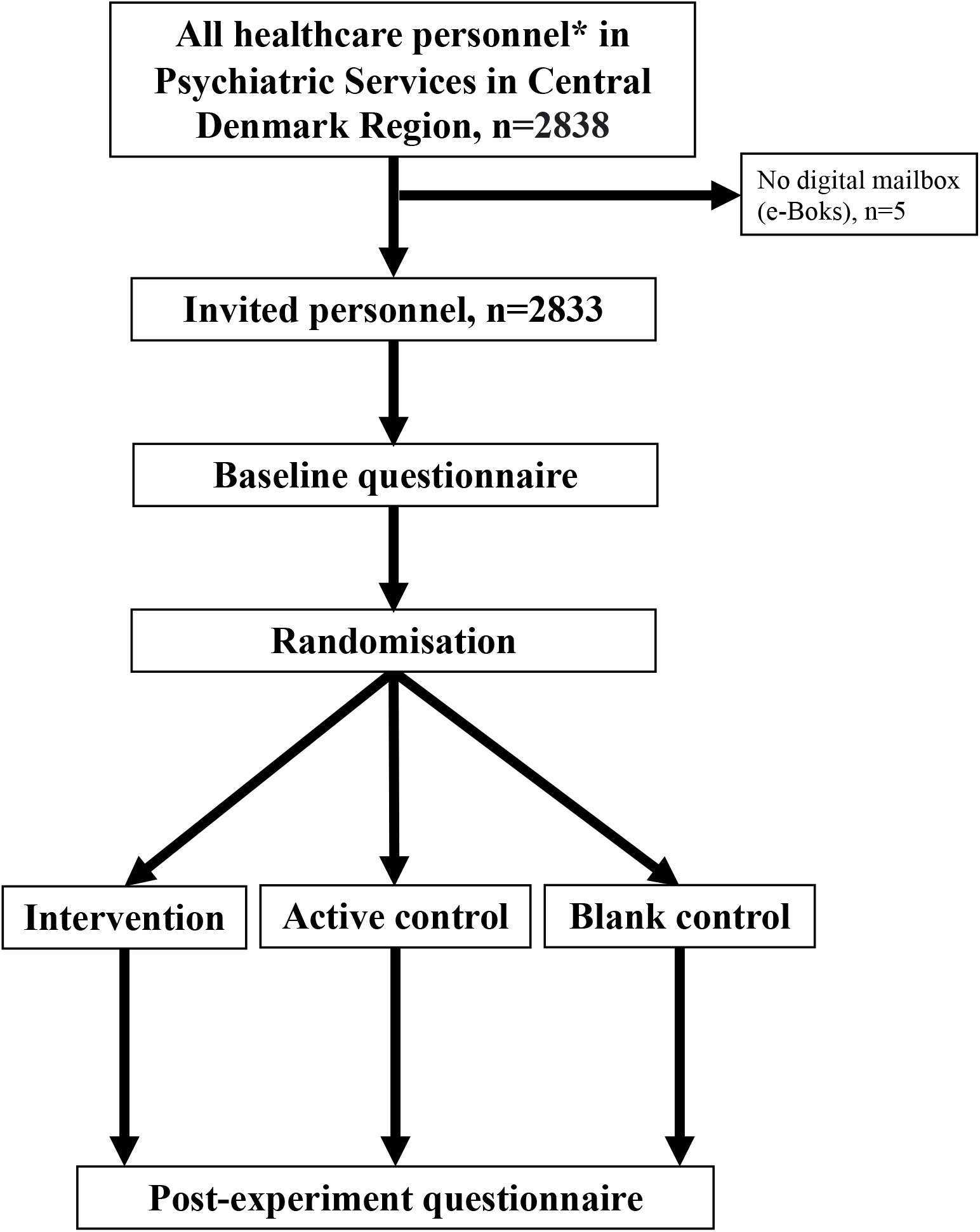
Flowchart of study design and population. *The following professions were invited: Clinical Managers, Doctor, Nurse, Psychologists, Physio-/Ergotherapists, Pedagogues, Social- and healthcare worker.e-Boks: The secure digital mailing system used by Danish authorities to communicate with citizens

### Setting

The survey experiment was conducted within the Psychiatric Services of the Central Denmark Region, covering a catchment area of approximately 1.3 million people, including urban and rural areas. The Psychiatric Services comprise five public psychiatric hospitals offering tax-financed inpatient, outpatient, and emergency psychiatric treatment.

### Participants

All doctors, nurses, psychologists, physiotherapists, ergotherapists and pedagogues employed full-time or part-time in the Psychiatric Services of the Central Denmark Region (n=2838) were invited to participate in the survey on February 5, 2024. The staff members received the invitation to participate via “eBoks” – the secure digital mailing system used by the Danish authorities to communicate with citizens.^6^ The participants provided consent by checking a box and accessed the survey via a link to the SurveyXact online survey service. Those not having participated by February 19, 2024 received a reminder via eBoks. Five staff members did not have eBoks and were not invited. No monetary incentive for participation was provided.

### Power calculation

In the analogue survey experiment among patients, 992 participants were sufficient to establish a statistically significant difference between the three randomisation groups.^5^ Assuming a similar effect size and variance among staff, we aimed for a similar sample size for the present study.

### Randomization

The participants were allocated to one of three arms: intervention, active control, and blank control (for a description, see the “Survey experiment” section, below). At the time of the study, SurveyXact did not support the standard method of random allocation (1/3, 1/3, and 1/3). Therefore, participants were allocated to one of the three groups based on the time they accessed the survey link provided in the invitation. Specifically, those who accessed the link between 0 and 0.333 seconds were allocated to the blank control group, those accessing between 0.334 and 0.666 seconds were allocated to the active control group, and those who accessed between 0.667 and <1 second were allocated to the intervention group. The participants were not informed about the randomization nor the experiment.

### Baseline questionnaire

The participants first completed a baseline questionnaire, irrespective of the allocation arm. The baseline questionnaire (available as Supplementary Table 1) assessed their current work status, workplace within the Psychiatric Services, marital status, household composition (adults and children), general trust, trust in technology, institutional trust, perceived understanding of ML-based clinical decision support systems, and experience with ML-based clinical decision support systems. Likert scales from 0 to 10 were used for the items regarding trust and perceived understanding.

### Experiment

After the baseline questionnaire, the participants received three different types of information according to the randomized allocation:

1. Intervention: An information pamphlet featuring both illustrations and text (slides within the electronic survey – see Supplementary Table 2), describing how an ML-based clinical decision support system works and may aid clinical practice in psychiatric services.
2. Active control: An information pamphlet featuring both illustrations and text (slides within the electronic survey – see Supplementary Table 3), describing a standard clinical decision-making process in psychiatric services without use of an ML-based clinical decision support system.
3. Blank control: No information pamphlet.

### Post-experiment questionnaire (outcome measure)

Following the survey experiment, the participants completed a questionnaire designed to assess their trust and distrust in ML-based clinical decision support systems in psychiatric services. Specifically, they answered the following questions: 1. “I feel safe that mental health professionals can make decisions with the support of machine learning models.” 2. “I trust that the Psychiatric Services can use machine learning models in a safe and appropriate way.” 3. “I am concerned that use of machine learning models for decision support in psychiatry will increase the risk of error.” 4.” I think that patients should have the opportunity to opt out of machine learning models being used for decision support in relation to their treatment in the psychiatric services.” 5. “I am concerned that healthcare services, including the psychiatric services, are becoming too dependent on machine learning models.” 6. “I am concerned that the use of machine learning models may lead to increased inequality in healthcare, including psychiatry.” 7. “The advantages of using machine learning models for decision support in psychiatry outweigh the disadvantages.” 8. “It is important to me that I can get an explanation of the basis on which a machine learning model recommends a given treatment”. 9. “I am concerned that a machine learning model may make incorrect recommendations due to inaccuracies in the medical record.” All questions were identical to those used in our survey experiment among patients^5^ except a slightly modification of item 4 and 9 as the original questions were intended for patients undergoing treatment (phrased as follows: Item 4:”I would like to have the opportunity to opt out of machine learning models being used for decision support in relation to my treatment in the psychiatric services.”; Item 9: “I am concerned that a machine learning model may make incorrect recommendations due to inaccuracies in my medical record.”). All responses were provided on an 11-level Likert scale ranging from 0 (“Totally disagree”) to 10 (“Totally agree”). At the end of the survey, the participants answered the exact same question regarding “perceived understanding of ML-based clinical decision support systems” as they did at baseline.

### Choice of primary outcome measure

As it is psychometrically suboptimal to aggregate positively and negatively worded items (following inversion),^7^ the three positively-worded “trust” items (1, 2, and 7) and the 5 negatively-worded “distrust” items (3, 4, 5, 6, and 9) were grouped a priori (see the pre-registered analysis plan). Item 8 was considered to be neutral and kept separate. Principal component analyses were conducted to assess if the trust and distrust items, respectively, loaded onto latent components. The number of components was determined by analysing the scree plot and selecting the number of components before the clear break (“elbow”) in the plot.^8^ Subsequently, an item was deemed to load onto a component if its loading was >0.40 or <-0.40.^9^ Based on the number of items loading onto each component, a trust total score and a distrust total score (the sum of the positively and negatively framed items, respectively) were constructed to serve as outcome measures.

### Handling of survey responses

Upon accessing the link to the survey, the participants entered their social security number. In cases where participants submitted the questionnaire multiple times, only their initial response was considered. If a participant initially provided a partial response, including the randomization element, and subsequently completed the survey, under a different randomization arm, their full response was excluded from analysis as they had become unblinded to the randomization. Since the questions serving as outcome measures were placed at the end of the survey, only completers were included in the analyses. Participants were required to answer all questions (no missing values).

### Statistics

The sociodemographic were summarized using descriptive statistics. Differences in survey completion time between the randomization groups were assessed using the Mann-Whitney U test. In the primary analyses, trust/distrust levels in ML-based clinical decision support systems, quantified by the trust/distrust scores, were compared pairwise between the three randomization arms via two-sample t-tests. Equivalent secondary analyses were performed for each individual trust/distrust item. Pearson’s correlation coefficient was used to evaluate the correlation between the trust and distrust scores. As part of the robustness analyses, two-sample t-tests of trust/distrust in ML-based clinical decision support systems was performed across the three randomisation arms, stratifying by sex, age (median split: <47 vs ≥ 47 years), profession (requiring master-level education vs. requiring bachelor-level education or less), baseline knowledge of machine learning as decision support (median split: <5 vs. ≥ 5), and the level of general trust (median split: <8 vs. ≥8). The significance threshold was set to 0.05. Correction for multiple comparisons was not conducted as the outcomes were highly interdependent.^10^ All data management and statistical analyses were performed using Rstudio version 2023.06.0 Build 421.

### Ethics

Research studies based on surveys are exempt from ethical review board approval in Denmark (waiver no. 1-10-72-109-23 from the Central Denmark Region Committee on Health Research Ethics). The study was approved by the Chief Medical Officer of the Psychiatric Services in the Central Denmark Region and registered on the internal list of research projects having the Central Denmark Region as data steward (reg. no. 1-16-02-12-24).

## Results

A total of 780 (27.5%) of the invited participants completed the online survey (100 partial respondents were excluded and 4 were excluded due to being unblinded to the randomization). Table 1 lists the sociodemographic characteristics of the participants and information regarding baseline knowledge about machine learning as a clinical decision support tool, general trust and trust in institutions.

**Table 1.** Characteristics of the 780 participants with complete responses.

| Variable | Overall<br>n = 780 | Randomization group |  |  |
| --- | --- | --- | --- | --- |
|  |  | Blank<br>control<br>n = 246 | Active<br>control<br>n = 252 | Intervention<br>n = 282 |
| **Age, median (IQR)** | 47 (22) | 46 (22) | 47 (22) | 47 (21) |
| **Sex, n (%)** |  |  |  |  |
| Female | 628 (81%) | 204 (83%) | 198 (79%) | 226 (80%) |
| **Profession, n (%)** |  |  |  |  |
| Clinical Manager | 19 (2%) | 8 (3%) | 5 (2%) | 6 (2%) |
| Consultant doctor | 46 (6%) | 15 (6 %) | 12 (5%) | 19 (7%) |
| Resident doctor | 63 (8%) | 20 (8%) | 21 (8%) | 22 (8%) |
| Nurse | 281 (36%) | 103 (42%) | 82 (33%) | 96 (34%) |
| Psychologist | 124 (16%) | 40 (16%) | 40 (16%) | 44 (16%) |
| Pedagogue | 65 (8%) | 15 (6%) | 21 (8%) | 29 (10%) |
| Physio-/Ergotherapist | 51 (7%) | 13 (5%) | 20 (8%) | 18 (6%) |
| Social and healthcare worker | 131 (17%) | 32 (13%) | 51 (20%) | 48 (17%) |
| **Employment status, n (%)** |  |  |  |  |
| Full-time employed | 543 (70%) | 177 (72%) | 176 (70%) | 190 (67%) |
| Part-time employed | 161 (21%) | 45 (18%) | 50 (20%) | 66 (23%) |
| Hourly pay/Substitute | 46 (6%) | 16 (7%) | 15 (6%) | 15 (5%) |
| Student worker | <5 (<2%) | <5 (<2%) | <5 (<2%) | <5 (<2%) |
| Sick leave | 15 (2%) | 5 (2.0%) | 7 (3%) | <5 (<2%) |
| Subsidized employment | 10 (1.3%) | <5 (<2%) | <5 (<2%) | 5 (1.8%) |
| Other | <5 (<2%) | <5 (<2%) | <5 (<2%) | <5 (<2%) |
| **Workplace, n (%)** |  |  |  |  |
| Aarhus - Department of Affective disorders | 113 (14%) | 40 (16%) | 30 (12%) | 43 (15%) |
| Aarhus - Department of Child- and adolescent psychiatry | 94 (12%) | 23 (9%) | 29 (12%) | 42 (15%) |
| Aarhus - Department of Forensic psychiatry | 75 (10%) | 15 (6%) | 29 (12%) | 31 (11%) |
| Aarhus - Department of Psychosis | 101 (13%) | 31 (13%) | 35 (14%) | 35 (12%) |
| Gødstrup - Department of Child- and adolescent psychiatry | 16 (2%) | 5 (2%) | <5 (<2%) | 8 (3%) |
| Gødstrup (incl. Holstebro)- Regional Psychiatry | 87 (11%) | 23 (9%) | 34 (13%) | 30 (11%) |
| Horsens -Regional Psychiatry | 61 (8%) | 26 (11%) | 17 (7%) | 18 (6%) |
| Randers (incl. Rønde) - Regional Psychiatry | 70 (9%) | 21 (9%) | 24 (10%) | 25 (9%) |
| Viborg - Department of Child- and adolescent psychiatry | 14 (2%) | 6 (2%) | <5 (<2%) | <5 (<2%) |
| Viborg (incl. Skive, Silkeborg) - Regional Psychiatry | 124 (16%) | 47 (19%) | 40 (16%) | 37 (13%) |
| Other | 25 (3%) | 9 (4%) | 7 (3%) | 9 (3%) |
| **Marital status** |  |  |  |  |
| Married | 432 (55%) | 136 (55%) | 148 (59%) | 148 (52%) |
| Domestic partnership | 131 (17%) | 42 (17%) | 36 (14%) | 53 (19%) |
| Non-domestic partnership | 81 (10%) | 26 (11%) | 29 (12%) | 26 (9%) |
| Divorced | 50 (6%) | 14 (6%) | 16 (6%) | 20 (7%) |
| Single | 86 (11%) | 28 (11%) | 23 (9%) | 35 (12%) |
| **Household adults, n (%)** |  |  |  |  |
| 1 | 167 (21%) | 48 (20%) | 51 (20%) | 68 (24%) |
| 2 | 431 (55%) | 135 (55%) | 140 (56%) | 156 (55%) |
| 3 | 148 (19%) | 50 (20%) | 51 (20%) | 47 (17%) |
| 4 or more | 34 (4.4%) | 13 (5.3%) | 10 (4.0%) | 11 (3.9%) |
| **Household children, n (%)** |  |  |  |  |
| 0 | 406 (52%) | 131 (53%) | 138 (55%) | 137 (49%) |
| 1 | 118 (15%) | 30 (12%) | 40 (16%) | 48 (17%) |
| 2 | 190 (24%) | 67 (27%) | 51 (20%) | 72 (26%) |
| 3 or more | 66 (8.5%) | 18 (7.3%) | 23 (9.1%) | 25 (8.9%) |
| **Baseline knowledge of machine learning, median (IQR)** | 5 (5) | 5 (5) | 5 (4) | 5 (5) |
| **Previously used a clinical decision support tool based on machine learning, n (%)** |  |  |  |  |
| Yes | 57 (7%) | 15 (6%) | 20 (8%) | 22 (8%) |
| No | 590 (76%) | 191 (78%) | 194 (77%) | 205 (73%) |
| I do not know | 133 (17%) | 40 (16%) | 38 (15%) | 55 (20%) |
| **General trust, median (IQR)** | 8 (2) | 8 (3) | 8 (2) | 8 (2) |
| **Trust in technology, median (IQR)** | 7 (2) | 7 (2) | 7 (3) | 7 (2) |
| **Trust in parliament, median (IQR)** | 6 (2) | 6 (2) | 6 (2) | 6 (3) |
| **Trust in judicial system, median (IQR)** | 8 (2) | 8 (2) | 8 (2) | 8 (2) |
| **Trust in the police, median (IQR)** | 8 (2) | 8 (2) | 8 (2) | 8 (2) |
| **Trust in the European Union, median (IQR)** | 6 (2) | 6 (2) | 6 (2) | 6 (2) |
| **Trust in healthcare services, median (IQR)** | 7 (2) | 7 (2) | 7 (2) | 7 (2) |
| **Trust in psychiatric services, median (IQR)** | 7 (2) | 7 (2) | 7 (2) | 7 (2) |

Cell counts <5 are not specified due to risk of identification of individual participants.

The participants were randomized with the following distribution: blank control = 246; active control = 252; intervention= 282. The response time had a median of 286 seconds (IQR = 150 seconds) for those assigned to the blank control arm, 362 seconds (IQR = 196 seconds) for the active control arm and, 389 seconds (IQR = 234 seconds) for the intervention arm. The active control- and intervention arm had a statistically significant (both p<0.001) longer response time than those in blank control but did not differ statistically significantly from each other (p-value= 0.23).

The findings from the principal component analysis are presented in Supplementary Tables 4-5 and Supplementary Figures 1-2. Consistent with a priori expectations, the survey items were categorized into two components: a trust component and a distrust component. As expected, the trust component consisted of the three positively worded items, but the distrust component only comprised of four negatively worded items (Outcome item 4: “Possibility to opt out” did not load sufficiently on the latent distrust component and was excluded). The sum scores for trust and distrust were strongly inversely correlated (Pearson correlation coefficient: -0.66).

Table 2 lists the responses to the items addressing trust and distrust in ML-based clinical decision support systems across the three randomization groups. Interestingly, the median scores for the trust items were generally within range with the baseline levels of general/institutional trust (see Table 1). Notably, the neutral “Explainability of ML models” item had the highest median score (10, interquartile range: 1).

**Table 2.**
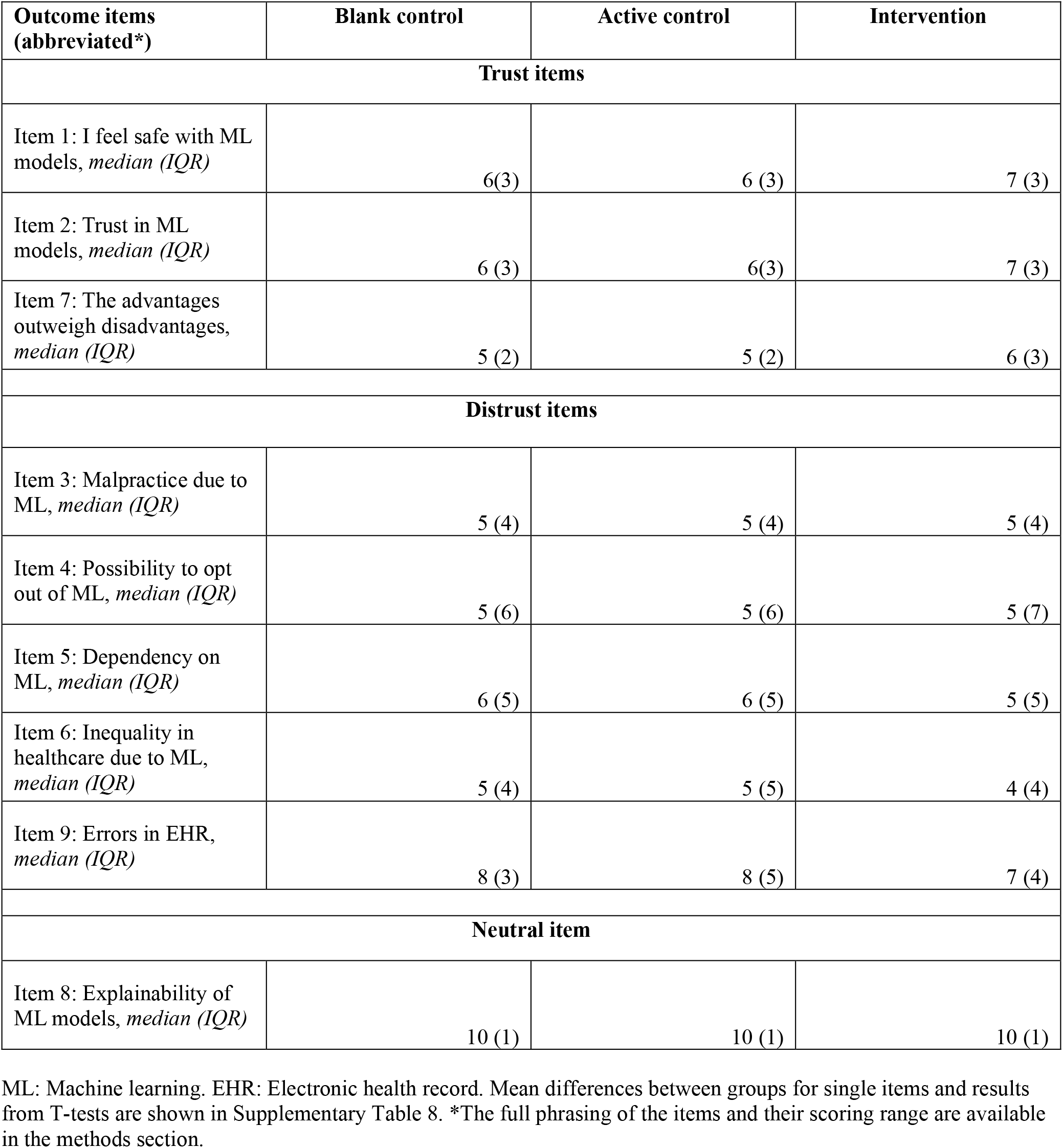
Individual item scores after the experiment.

Figure 2 illustrates the results of the primary analysis, comparing the three randomization groups on trust and distrust sum scores. The intervention led to an increase in trust in ML-based clinical decision support systems compared to both the active control (mean difference in trust: 5% [95% CI: 2%, 9%], p<0.001) and the blank control (mean difference in trust: 5% [2%, 11%], p=0.003). Similarly, the intervention decreased distrust compared to the active control (mean difference in distrust: -4% [-7%, -1%], p=0.004) and the blank control (mean difference in distrust: -3% [-6%, - 1%], p=0.021). There were no significant differences in trust or distrust between the active and blank control groups.

**Figure 2.**
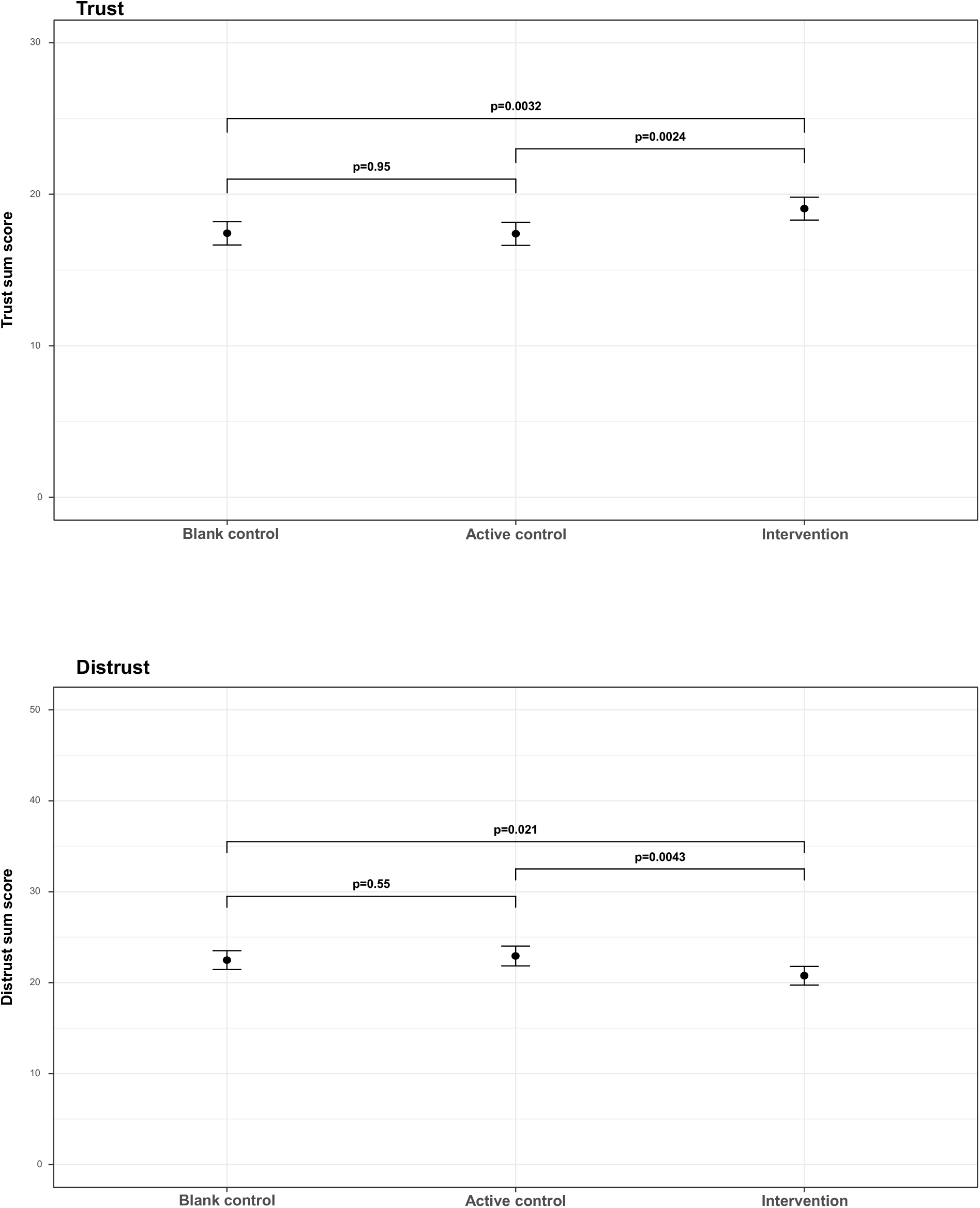
Effect of the intervention on trust (top) and distrust (bottom) in machine learning model-based clinical decision support systems. The error bars represent confidence intervals.

Results of the comparisons between the randomization groups on the individual trust and distrust items (align with the sum score analyses) are available in Supplementary Table 6. For the neutral item “Explainability of ML models” item, there were no statistically significant differences between the randomization groups.

Results of the analyses of the intervention effect stratified by sex, age, profession, baseline knowledge of machine learning as decision support, and the level of general trust are listed in Supplementary Table 7. Generally, the intervention effect remains consistent across strata, with a few notable exceptions. Specifically, the intervention increased trust and decreased distrust in the older half of the population, but not/less in the younger subgroup. Similarly, the intervention increased trust and decreased distrust those with a profession “Requiring bachelor-level education or less”, but not/less in those with a profession “Requiring master-level education”.

The intervention group exhibited a statistically significantly greater pre-post experiment increase in perceived understanding of ML-based clinical decision support systems compared to both the active control (mean difference: 4.27% [95% CI: 2.64%, 5.89%], p<0.001) and the blank control (mean difference: 5.50% [95% CI: 3.93%, 7.08%], p<0.001). There was no statistically significant difference between the blank control and active control groups.

## Discussion

Through an online randomized survey experiment among healthcare staff employed in a psychiatric service system (that of the Central Denmark Region), we showed that receiving information regarding ML models as clinical decision support systems increased trust and decreased distrust in them. Notably, the findings also revealed that explainability of ML models in the context of clinical decision support held particular importance for the participating staff.

To the best of our knowledge, this is the first study to examine if receiving information on machine learning as clinical decision support systems can modify healthcare staff’s trust and distrust in this technology. The results align almost 1:1 with the observations from our analogue survey experiment (same intervention and same outcome assessment) conducted among patients receiving treatment in the Psychiatric Services of the Central Denmark Region.^5^ Taken together, these findings suggest that the effect of receiving basic information about machine learning as a clinical decision support system generalizes across patients and staff.

The difference in intervention effect between those with professions requiring bachelor-level education and those with professions requiring master-level education detected in this study is likely attributed to the lower baseline self-reported knowledge of ML among the former (mean difference [95% CI] = 0.63 [0.194;1.08]), which may have contributed to a ceiling effect (less room for increasing trust/reducing distrust among those with higher education and self-reported baseline knowledge of ML). The intervention also had a larger effect in the older fraction of the staff compared to the younger fraction. Although, there was no substantial difference in self-reported baseline knowledge of ML between these groups (mean difference [95% CI] = 0.13[-0.27;0.54]), prior research^11^ describes age differences in knowledge of ML in the general population, which points to a potential ceiling effect for the younger age group. Taken together, these results suggest that education on ML as clinical decision support systems would likely benefit from being tailored to the specific target group at hand – and that characteristics such as educational background and age should be factored into this tailoring.

That explainability of ML models was deemed particularly important by the participating staff aligns with findings from studies among healthcare staff in general,^4^ among various patient populations (e.g., within primary care, radiology, dermatology, and our own findings from psychiatry),^5,12,13^ and the general population.^12,14^ This is likely one of the reasons for the strong emphasis on explainability in the recent AI Act from the European Union (EU).^15,16^ Specifically, under “Human Oversight” (article 14), the EU AI act states that “…natural persons to whom human oversight is assigned are enabled, as appropriate and proportionate: … to correctly interpret the high-risk AI system’s output, taking into account, for example, the interpretation tools and methods available.” This refers both to “local explanations”, i.e., the interpretation and explainability of a specific model, as well as “global explanations”, i.e., that users understand the overall function of high-risk machine learning/AI systems. ^16^

That healthcare professionals in general will likely benefit from being educated with regard to artificial intelligence (AI)/ML has been proposed elsewhere. In a survey of medical students from Germany in 2019, two-thirds of the respondents did not consider themselves to have baseline knowledge about AI/ML (this number has likely decreased since then due to the launch and availability of AI-driven chatbots such as ChatGPT), and 71% agreed that AI/machine learning should be included in their curriculum.^**17**^ Furthermore, the American Medical Association has issued policy recommendations emphasizing that education is a crucial component of the implementation of AI in medicine.^**18**^ Our findings corroborate these views and indicate that information is indeed helpful in increasing understanding of these systems, suggesting that educating healthcare staff is essential for the future of medical machine learning/AI.

There are limitations to this study, which should be considered when interpreting the results. First, the study had a response rate of 27.5%, which could have resulted in selection bias. Due to the lack of sociodemographic data on those not participating, we were unable to adjust the analyses for attrition. However, when compared to similar survey studies among healthcare staff, the response rate of the present study is relatively high.^13,19^ Second, we did not employ a validated questionnaire to measure trust and distrust in machine learning (ML) as a clinical decision support tool, as such a tool is, to our knowledge, not yet available. Despite this limitation, we believe that the underlying components of trust and distrust, identified through principal component analysis of items with apparent face validity, serve as reasonable outcome measures. Third, we did not include attention check questions, and some participants may, therefore, have responded inconsistently or arbitrarily, adding noise to the data - complicating signal detection. Nevertheless, a signal was observed, indicating that the intervention increased trust and reduced distrust. Fourth, while the randomized and controlled design of this study suggests that the intervention had a causal effect on trust and distrust, we have not investigated the longevity of it. Additionally, the observed effect size was numerically small, which was anticipated given the complexity of trust as a construct,^20^ and the intervention comprising only 4 pages of visual and textual information. In future studies, efforts should be made to extend the post-intervention observation period and exploring the effect of repeated or more intensive interventions. Fifth, while the results of this study highlight the potential of educating staff from psychiatric services with regard to ML models as clinical decision support systems, it can be argued that both the intervention (information on ML models in psychiatry in general) and the outcome (general trust/distrust in ML models) were too generic. Therefore, future research would benefit from focusing on more specific ML models as well as the precise level of information required to achieve well-calibrated trust/distrust – as blind trust is of course not the goal. Sixth, with regard to generalizability, it is important to bear in mind that Denmark is one of the most digitalized countries in the world, ^21^ and its population may be more receptive to new technology. Therefore, replicating the results of the present study in other countries would be ideal. We have translated the survey to English (available in the Supplementary Material) to enable replication.

In conclusion, this survey experiment indicates that providing information on ML-based clinical decision support systems to healthcare staff members in psychiatric services increases their trust in such systems. This finding aligns with those from studies of other healthcare professionals, patient populations, and the general population, suggesting that offering appropriate information is a vital part for a successful implementation of ML-based clinical decision support systems.

## Data Availability

The data cannot be shared as the participants have not consented to data sharing.

## Contributors

All authors contributed to the conceptualization and design of the study. EP and SDØ conducted and supervised the data collection. Data management and verification was performed by EP. Statistical analysis was carried out by EP. All authors contributed to the interpretation of the obtained results. EP wrote the first draft of the manuscript, which was subsequently revised for important intellectual content by the other authors. All authors approved the final version of the manuscript prior to submission.

## Acknowledgments

There was no specific funding for this study. Outside this study, SDØ reports funding from the Lundbeck Foundation (grants R358-2020-2341 and R344-2020-1073), the Novo Nordisk Foundation (grant NNF20SA0062874), the Danish Cancer Society (grant R283-A16461), the Central Denmark Region Fund for Strengthening of Health Science (grant 1-36-72-4-20), the Danish Agency for Digitisation Investment Fund for New Technologies (grant 2020-6720), and Independent Research Fund Denmark (grant 7016-00048B and 2096-00055A). These funders played no role in the design or conduct of the study; collection, management, analysis, and interpretation of the data; preparation, review, or approval of the manuscript; and decision to submit the manuscript for publication.

The authors are grateful to Henrik Gersfelt from the Central Denmark Region for data management, and to Torben Schmidt Kjeldsen from the Central Denmark Region for graphical design of the survey experiment.

## Data sharing

The data cannot be shared as the participants have not consented to data sharing.

